# Monkeypox outbreak in the Netherlands in 2022: public health response, epidemiological and clinical characteristics of the first 1000 cases and protection of the first-generation smallpox vaccine

**DOI:** 10.1101/2022.10.20.22281284

**Authors:** Catharina E. van Ewijk, Fuminari Miura, Gini van Rijckevorsel, Henry J.C. de Vries, Matthijs R.A. Welkers, Oda E. van den Berg, Ingrid H.M. Friesema, Patrick van den Berg, Thomas Dalhuisen, Jacco Wallinga, Diederik Brandwagt, Brigitte A.G.L. van Cleef, Harry Vennema, Bettie Voordouw, Marion Koopmans, Annemiek A. van der Eijk, Corien M. Swaan, Margreet J.M. te Wierik, Tjalling Leenstra, Eline Op de Coul, Eelco Franz, the Dutch Monkeypox Response Team

## Abstract

In early May 2022 a global outbreak of monkeypox (MPX) started among persons without a travel history to regions known to be enzootic for monkeypox-virus. On August 8 2022, the Netherlands reported its 1000^th^ monkeypox case representing a cumulative incidence of 55 per million population, one of the highest cumulative incidences worldwide. Here we describe the epidemiological characteristics and clinical presentation of the first 1000 monkeypox cases in the Netherlands, within the context of the public health response. Additionally, we explored risk factors for and estimated the protective effect of first-generation smallpox vaccine against more severe MPX.

The first 1000 MPX cases, reported between May 20 and August 8 2022, were predominantly MSM aged 31-45 years. The vast majority of infections were acquired through sexual contact with casual partners in private or recreational settings including LGBTQIA+ venues in the Netherlands. This indicates that, although some larger upsurges occurred from point-source and/or travel related events, the outbreak is mainly characterised by sustained transmission within the Netherlands. More severe MPX was associated with having one or more comorbidities as well as having participated in more (3+) different sexual activities 21 days before symptom onset. We found a vaccine effectiveness of the prior first-generation smallpox vaccine against more severe MPX of 58% (95% CI 17-78%), suggesting moderate protection against more severe MPX symptoms on top of any possible protection by this vaccine against MPXV infection and disease.

## Introduction

In the first week of May 2022 the United Kingdom reported cases of monkeypox virus (MPXV) among persons without travel histories to regions known to be enzootic for MPXV such as West and Central Africa. ^1^ Within weeks, multiple cases were reported in Europe, North-America and Australia. ^2-4^ Unlike previous reported outbreaks of monkeypox (MPX) in the African region and the US, almost all cases were men who (also) have sex with men (MSM), particularly those with multiple sexual partners.

Immediately after the first MPX case was confirmed on May 20 in Amsterdam, the Dutch government declared monkeypox a notifiable group A disease (May 21) to ensure centralized coordination of the response and swift public health action, including mandatory notification of suspected and confirmed cases by treating physicians and laboratories within 24 hours, case isolation, and source and contact tracing. The Netherlands reported the 1000^th^ case on August 8 2022 representing a cumulative incidence of 55 per million population; one of the highest cumulative incidences worldwide, after Spain (104/1 million) and Portugal (69/1 million) at that time, and with 31,112 confirmed MPX cases reported worldwide. ^5^

As the Netherlands stopped the first-generation smallpox vaccination campaign in 1974 and infections with viruses from the genus Orthopox virus are rare, more than half (57%) of the Dutch population has never been exposed to Orthopox viruses and can be considered immunologically naive. ^6, 7^ Worldwide, the World Health Organization stopped the smallpox vaccination campaign in 1977 followed by declaring smallpox eradicated in 1980. ^8^ Individuals who have been vaccinated with the first-generation smallpox vaccine might, however, still benefit from some cross-protection against (severe) MPX. ^9^ Here we describe the epidemiological characteristics and clinical presentation of the first 1000 MPX cases in the Netherlands within the context of the public health response, and we provide estimates of the protection against more severe (i.e. non-mild) MPX symptoms offered by the first-generation smallpox vaccine.

## Methods

### Study design and population

An observational study of persons tested for Orthopoxvirus with MPXV confirmation or primary MPXV detection and reported to the Netherlands National Institute for Public Health and the Environment (RIVM) between May 20 and August 8 2022.

### Definitions

Cases were reported as confirmed, probable and possible cases (Supplement 1, Box 1). Confirmed MPX cases were categorised into mild or more severe MPX. Mild MPX was defined as cases experiencing either 1-2 systemic symptoms (e.g. fever, lymphadenopathy) and/or skin lesions on one body location (e.g. head, limps, trunk, peri-anal, genital). More severe MPX was defined as cases experiencing ≥3 systemic symptoms and/or skin lesions on ≥2 body locations and/or hospitalisation due to MPX. Country of origin was categorised according to the definition of Statistics Netherlands (the Netherlands, Turkey, Morocco, Netherlands Antilles, Surinam and Aruba, other Western country – including Europe, North-America, Oceania, Indonesia and Japan – and other non-Western countries– all other). ^10^ Persons with high-risk of MPXV exposure were defined as individuals who engaged in group sex, sex on premises (e.g. LGBTQIA+ sauna’s and cruising areas) in the Netherlands or abroad, or who were a contact of a confirmed MPX case 21 days before symptoms onset. Contacts were categorised in high-, medium- and low-risk (Supplement 1, Box 2).

### Data collection

Data were collected as part of routine surveillance based on obligatory notification of probable, suspected and confirmed cases. To ensure notification of MPX cases from the regional Public Health Services (PHS) to the RIVM, a questionnaire was included in the national surveillance system for notifiable diseases (OSIRIS) for data collection on clinical and epidemiological information on cases, including demographic data, symptoms, medical history, and potential source(s) of infection. Data on MPX cases reported to the RIVM from May 20 until August 8 2022 were extracted from the OSIRIS database for analyses.

### Laboratory methods

Laboratory confirmation of MPXV was performed on pharyngeal-, skin lesion(s)-, and/or anal swabs collected either into virus transport medium or on dry swabs or e-swabs. Samples were tested by real-time Polymerase Chain Reaction (RT-PCR). Diagnostic protocols were validated and based either on pan-Orthopox RT-PCR with subsequent MPXV detection through sequence analysis or had a monkeypox-specific target. ^11-13^

### Statistical analysis

Descriptive analysis was performed for demographics of MPX cases, their epidemiological characteristics including the most likely route -, source - and place of transmission.

The reporting delay (days) between symptom onset and notification as a confirmed MPX case was calculated. Based on the calculated reporting delay, we estimated the daily reported confirmed cases by date of symptom onset by correcting for underreporting (i.e. nowcasting). ^14^

Univariable and multivariable analyses (adjusting for age) by using logistic regression were performed to assess risk factors for more severe MPX.

Vaccine effectiveness (VE) of the first-generation smallpox vaccine against more severe MPX was calculated among individuals born before 1978 by comparing the odds of first-generation smallpox vaccination between persons with more severe compared to those with mild MPX. VE was calculated as 1 minus the odds ratio (OR). The crude OR and 95% confidence intervals (CI) were calculated using logistic regression analyses. Individuals who had received Imvanex®, the in the European Union approved third-generation smallpox vaccine, as post-exposure prophylaxis (PEP) (n=40) were excluded from VE analyses due to its likely influence on symptom development. Additionally, VE was estimated adjusting for age (44-50, 51-55, and 56+ years). Statistical analyses were conducted using R version 4.0.2.

### Ethics

This study received medical ethical clearance (EPI-584) by the Centre for Clinical Expertise (KEC) at the RIVM.

## Results

### Public Health Response

Due to the increasing number of MPX cases in neighbouring European countries in the first weeks of May 2022, a monkeypox response team (RT) was convened on 18 May at the Centre for Infectious Disease Control (CIb) of the RIVM, consisting of experts from the CIb, representatives of regional Public Health Services (PHS), the external reference laboratory (Erasmus Medical Centre (EMC)), the medical expert group on Sexual Health and Sexually transmitted Infections (WASS), Soa Aids Nederland foundation, and infectious disease specialists. The RT provided scientific advice (on request of the Ministry of Health, MoH) on diagnostics, risk assessment, risk classification and infection prevention and control measures regarding cases and contacts (including notifiable disease status and pre- and post-exposure vaccination). The RT also coordinated the communication for the at risk population, professionals and the general public. After the declaration of MPX as group A notifiable disease by the MoH, health care professionals were required to notify suspected MPX cases as soon as possible to their regional PHS for immediate public health actions (Supplement 1, Box 1). ^15^ After notification, a public health team contacted the case to advise self-isolation and hygiene measures, and to arrange diagnostic testing. Information on demographics including age, sex, sexual orientation, symptoms, date of symptom onset, hospitalisation, and details on source-(e.g. risk behaviour and potential sources of infection in the 21 days before symptom onset) and contact tracing (inventory numbers of high-, moderate-, and low-risk contacts) was collected. After MPX diagnosis was confirmed, high- and medium-risk contacts were informed and monitored by telephone for development of symptoms up to 21 days after last exposure. In addition, high-risk (and some medium-risk) contacts were offered the Imvanex® vaccine as PEP – preferably administered within 4 and up to 14 days after first exposure. At first, contacts who had sexual, intimate skin-to-skin or household contact (high-risk contact a, b and c in Box 2, Supplement 1) with MPX cases were asked to self-quarantine for 21 days. However, on June 24, on advice of a national council of experts (Deskundigenberaad), this quarantine measure was replaced with the advice to refrain from intimate (and sexual) contact during the 21-day monitoring period. This policy was adapted to overcome challenges experienced during source and contact tracing, such as hesitance by index cases to reveal contact information of their (sexual) contacts because of the potential consequences of prolonged quarantine. A second policy change was made on July 7, to offer Imvanex® as pre-exposure vaccination (PrEP) to MSM and transgender individuals who: 1) receive HIV-PrEP (or are on a HIV-PrEP waiting list) via Sexual Health Centers (SHC) or general practitioners (GPs), 2) are living with HIV and at increased risk of sexually transmitted infections (STI), 3) are known at the SHC or GP and at increased risk of STI, including MSM sex workers. ^16^ The vaccination campaign started on July 25 in the two public health regions with the highest MPX incidence (PHS of Amsterdam and The Hague), subsequently followed by all regions in the Netherlands.

### Communication

The RIVM provided information on MPX to the general public (e.g. on disease characteristics, symptoms and prevention) through the RIVM website and (social) media. Professionals (e.g. medical microbiologist, infectious disease physicians, SHC and PHS professionals) were regularly informed on e.g. clinical characteristics, (new) diagnostics and control measures, and the epidemiological situation through direct messaging services (Inf@ct and Signaleringsoverleg) and the publication of a MPX guideline. ^17-19^ Soa Aids Nederland – a Dutch STI policy and prevention foundation for professionals and the public – coordinated the MPX communication campaign targeting the MSM community. Communication materials such as posters, flyers, and digital content were developed to improve knowledge on and raising awareness for MPX (e.g. symptoms, partner notification, self-isolation, vaccination, and other risk-reducing measurements). These materials were distributed to all PHS, HIV treatment centres, and Lesbian Gay Bisexual Transgender Queer Intersex and Asexual+ (LGBTQIA+) venues across the country, including gay clubs, -saunas and other (sex) venues, and social events such as Gay Prides. LTBTQIA+ club owners and - event organisers were informed about MPX, transmission risks, and prevention measures (such as hygiene guidelines) for sex-on-premises. A manual was developed for PHS to guide outreach control measures and to start a dialogue about MPX with the MSM community. ^20^ Online campaigns included information on MPX (prevention) on websites, Facebook and Instagram accounts from organisations targeting LGBTQIA+ communities (such as <u>Man-tot-Man</u> from the PHS of Amsterdam, PrEP-NU, several regional PHS website, and COC Netherlands – an organisation advocating the rights of LGBTQIA+ people, as well as on gay-dating apps, such as Grindr® and Recon®). Information and real-life stories were shared through podcasts. Additionally, Soa Aids Nederland facilitated and provided financial support to the MSM community to organise their own MPX webinar (‘Het Grote Monkeypox Informatie Webinar’ 3 August 2022).

### Outbreak description

From the confirmation of the first MPX patient in the Netherlands on May 20 until August 8 2022 a total of 1,928 individuals were tested for MPXV, of whom 1,086 (56%) were initially defined as a probable and 842 (44%) as a possible case. In total 1000 (55%) individuals tested positive and 806 (45%) tested negative for MPXV. On August 8, results were pending for 122 persons, and therefore excluded from analyses. Test positivity among probable cases was 87% (902/1,040) and 13% (98/766) among possible cases.

The 1000 confirmed MPX cases had a symptom onset between April 27 and August 2, 2022 (Figure 1). Most cases (536/1000; 54%) were reported by the PHS of Amsterdam, followed by The Hague (85/1000; 8%) and Rotterdam-Rijnmond (60/1000; 6%). Within 1 week after detection of the first case in Amsterdam, 6 out of 25 (24%) PHS regions in the Netherlands had reported one or more MPX case(s), with 20 out of 25 (83%) within 1 month and all 25 PHS regions within 2 months.

**Figure 1.**
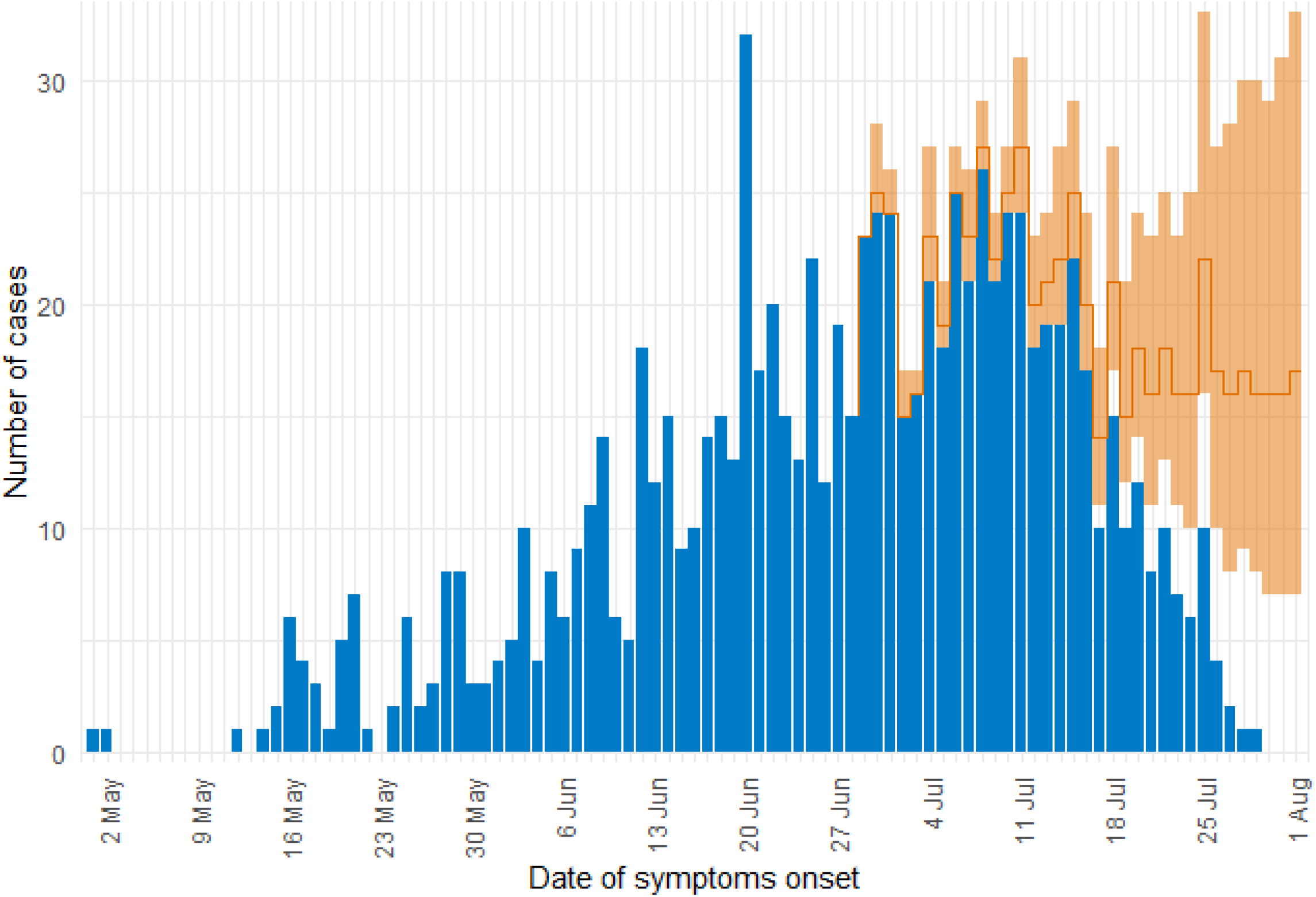
Laboratory confirmed monkeypox cases by date of symptom onset (N=894, blue bars), The Netherlands, May 20 to August 1 2022, and nowcast of confirmed cases of onset on August 1 (orange line), with 90% (shaded areas) prediction intervals.

The reporting delay, measured from time of symptom onset to reporting as a confirmed MPX case in OSIRIS was on average 12 days (median 10 days). The number of reported cases by date of symptom onset reached a peak in the first half of July 2022.

### Demographic and epidemiological characteristics

Of all MPX cases, 987/997 (99%) were men of whom 935/987 (95%) identified as MSM (gay, bisexual, or other MSM). Median age was 37 years (interquartile range: IQR 31-45, range 9-77). Fifty-seven percent (511/893) of the cases was born in the Netherlands, followed by 19% (173/893) in non-Western and 18% (159/893) in other Western countries.

Thirty-nine percent (343/882) reported one or more comorbidities, of which HIV-infection and other STIs were the most common. Medication use was reported in 476/811 (59%) of the cases, with HIV-PrEP (265/811; 33%) and HIV antiretroviral treatment (168/811; 21%) most frequently reported. In total, 227/678 (33%) had been a notified contact (predominantly high-risk contacts) of a monkeypox case, of whom 18% (40/227) received Imvanex® PEP. Thirteen percent (126/948) of the cases had received first-generation smallpox vaccine prior to this outbreak. A median of one high-risk and zero medium risk contacts was reported per case, but ranged from 0-100 and 0-99 contacts respectively (Table 1).

**Table 1.**
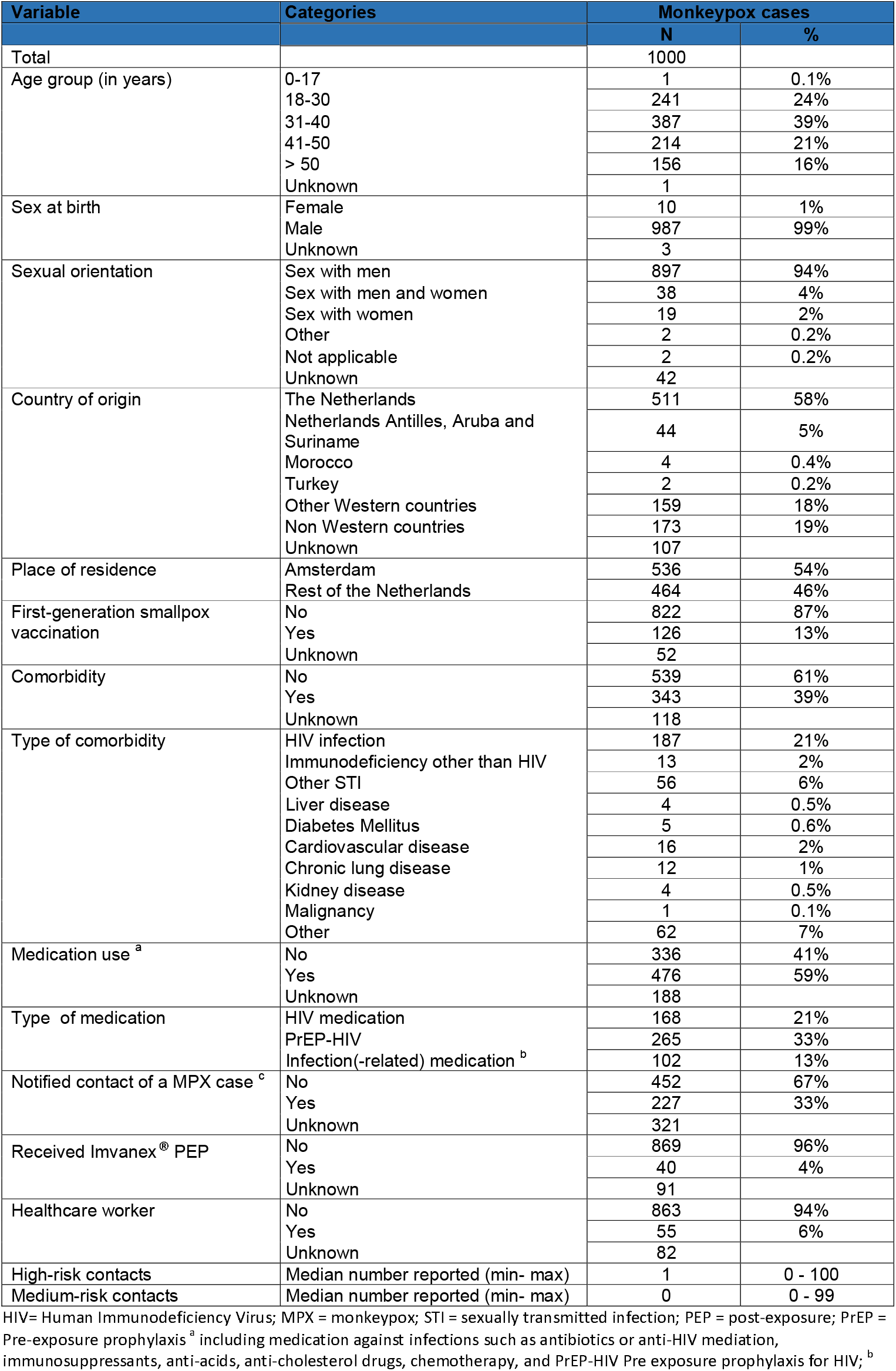
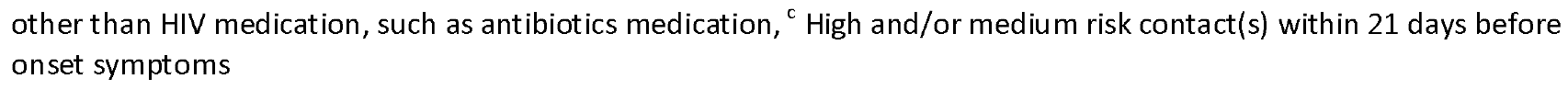
Demographic characteristics of laboratory confirmed monkeypox cases (N=1000) in the Netherlands, May 20 to August 8 2022.

Epidemiological data on cases identifying as heterosexual were often lacking. Of 19 male heterosexual cases, eight reported sexual contact as most probable transmission route, with 15 most likely in the family/home setting, and two were notified contacts of a MPX case. Of the 10 female cases, five reported sexual contact (with a MSM) as most probable transmission route, with eight most likely in the family/home setting, and five were notified contacts.

### Clinical characteristics

In total 850/991 (86%) MPX cases reported systemic symptoms (such as fever and lymphadenopathy), 914/991 (92%) reported skin lesions, and 796/968 (82%) both systemic symptoms and skin lesions. Fifty-four (6%) and 118 (12%) cases reported only systemic symptoms, and skin lesions respectively.

For half of the cases who experienced both skin lesions and systemic symptoms (316/632), the development of skin lesions was reported up to 9 days (median 2 days) after first systemic symptom(s) onset. In 24% of cases (154/632) the skin lesion(s) and systemic symptom(s) developed on the same day. In 26% of cases (162/632) the development of systemic symptoms were reported up to 13 days after the first skin lesion(s) (median 3 days).

Half of the cases (425/830, 51%) reported multiple (≥3) systemic symptoms, of which fever (521/991, 53%) and lymphadenopathy (371/991, 37%) most frequently. Only 43/991 (4%) reported coughing and 81/991 (8%) other respiratory symptoms. Most cases (573/912, 63%) reported lesions on 2-3 different body locations: mostly genital and/or perianal (75%), followed by genital only (51%), the limbs (51%) trunk (38%), and peri-anal only (33%). The lesions were mostly vesicular (536/991, 59%) and pustular (420/991, 46%). Twenty-eight percent (280/991) of cases had mild MPX and 72% (711/991) more severe MPX symptoms (Table 2). One infection was detected in a child and details have been published. ^21^

**Table 2.**
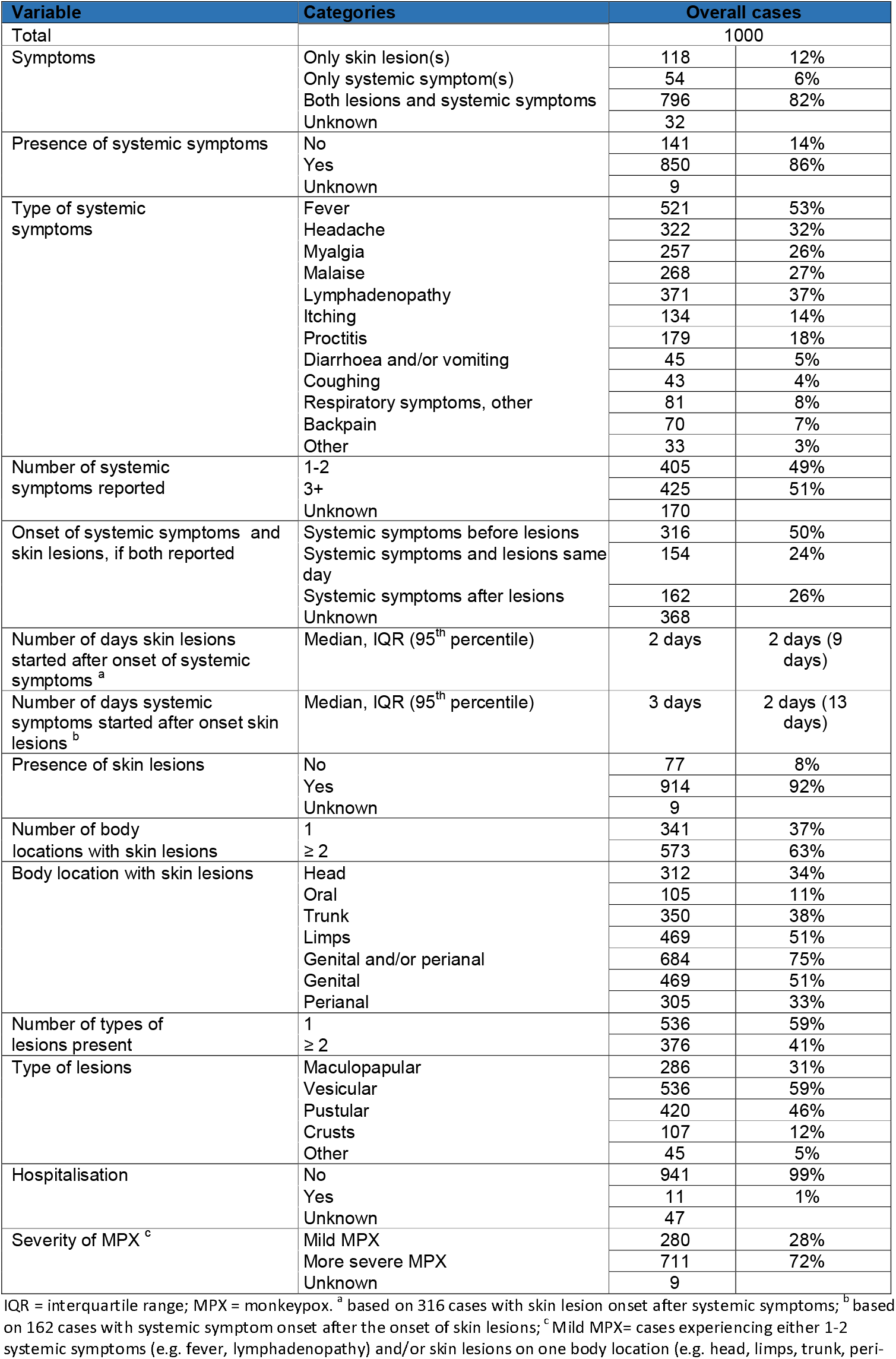
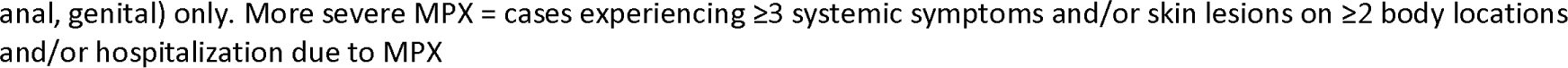
Clinical characteristics of monkeypox cases (N=1000), the Netherlands, May 20 to August 8 2022.

### Transmission routes and exposure

Sexual contact (including mucosal and skin-skin contact) was the dominant reported route of transmission (822/865; 95%). Transmission most likely occurred in a family/home setting (417/977, 43%), other recreational settings such as sports clubs or house parties (217/977, 22%), and LGBTQIA+ nightlife in the Netherlands (201/977, 21%). Most cases (577/920; 63%) had not travelled abroad in the 21 days before symptom onset, contrary to the beginning of the outbreak when 26/39 (67%) cases in May reported travel compared to 192/526 (37%) in July and 15/53 (28%) in August. Of those who travelled, European countries were mostly visited (316/343, 92%), predominantly Germany (n=99), Spain (n=94), and Belgium (n=44). Sixteen percent (150/) of cases reported participation in sexual activity whilst abroad. Most cases (539/861, 63%) had not visited (nightlife/entertainment/meeting) venues in the Netherlands 21 days prior to symptom onset. For those who did, LGBTQIA+ sex venues/meeting places (144/861, 17%) were mostly visited, followed by other nightlife/entertainment venues (117/861, 14%). Twenty-three percent (194/861) reported participation in sex on premises, and 44% (315/708) in group sex 21 days prior to symptom onset. Sexual partners were mostly casual (708/771, 92%).

### Risk factors for more severe monkeypox

Comparing more severe to mild MPX cases and adjusting for age, two out of eleven risk factors showed a (borderline) significant positive association with more severe MPX: suffering from one or more comorbidities (OR 1.5; 95% CI 1.1 – 2.1), and participating in 3+ sexual activities (such as oral, anal, and/or oral-anal sex) compared to 1-2 sexual activities in the 21 days before symptom onset (OR1.5, 95%CI 1.0 – 2.1) (Table 4).

**Table 3.**
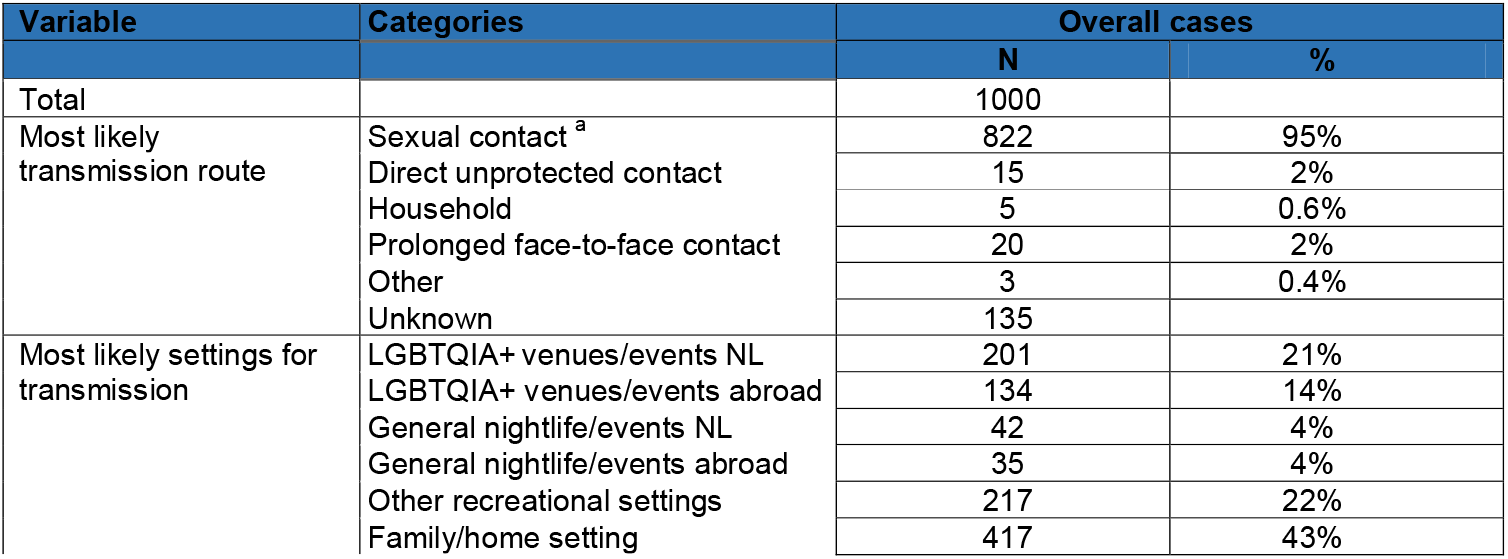
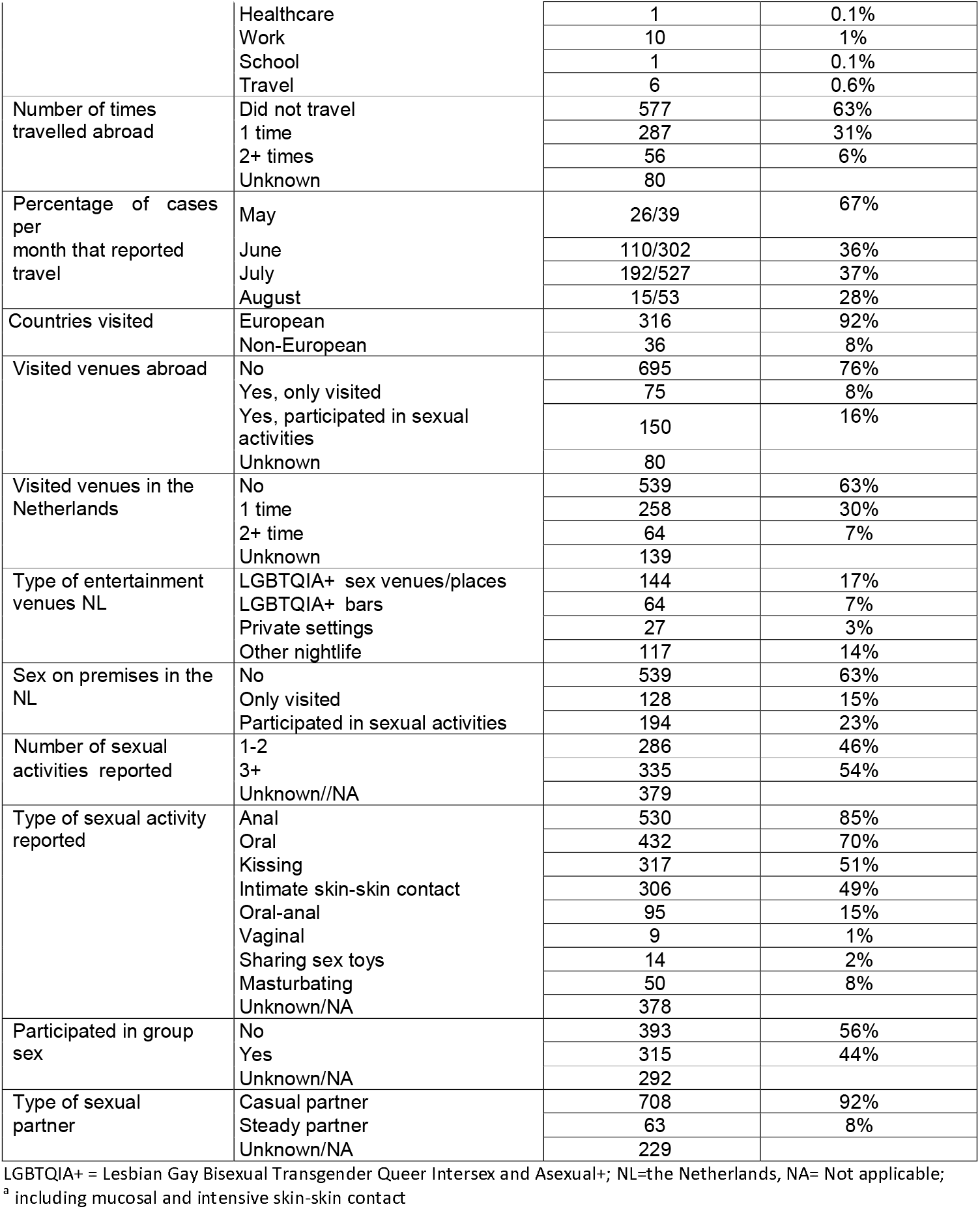
Reported transmission routes of infection and behavioural characteristics of monkeypox cases (N=1000), the Netherlands, May 20 to August 8 2022.

**Table 4.** Risk factors for more severe compared to mild monkeypox disease ^a^, the Netherlands, May 20 to August 8 2022.

| variable |  | Unadjusted odds ratio | 95% CI | Adjusted <sup>b</sup> odds ratio | 95% CI |
| --- | --- | --- | --- | --- | --- |
| **Age in years** | 0-30 | Ref. |  |  |  |
|  | 31-40 | 1.1 | 0.7 – 1.6 |  |  |
|  | 41-50 | 0.6 | 0.4 – 0.9 |  |  |
|  | 51+ | 0.4 | 0.3 – 0.6 |  |  |
| **Sex at birth** | Female | Ref. |  |  |  |
|  | Male | 1.7 | 0.4 – 6.0 |  |  |
| **Sexual orientation** | No MSM | Ref. |  |  |  |
|  | MSM | 1.5 | 0.6 – 3.4 |  |  |
| **Country of origin** | The Netherlands | Ref. |  | Ref. |  |
|  | Other countries | 1.1 | 0.8 – 1.5 | 1.6 | 0.6 - 3.8 |
| **Comorbidity** | No | Ref. |  | Ref. |  |
|  | Yes | 1.2 | 0.9 – 1.6 | **1.5** | **1.1 - 2.1** |
| **HIV infection** | No | Ref. |  | Ref. |  |
|  | Yes | 0.9 | 0.7 – 1.4 | 1.1 | 0.8 - 1.7 |
| **Medication <sup>c</sup>** | No | Ref. |  | Ref. |  |
|  | Yes | 0.9 | 0.7 – 1.3 | 1.0 | 0.7 - 1.4 |
| **Imvanex<sup>®</sup> PEP** | No | Ref. |  | Ref. |  |
|  | Yes | 0.6 | 0.3 – 1.3 | 0.6 | 0.3 - 1.2 |
| **Contact of MPX case** | No | Ref. |  | Ref. |  |
|  | Yes | 1.3 | 0.9 – 1.8 | 1.2 | 0.8 - 1.8 |
| **Transmission route** | Other | Ref. |  | Ref. |  |
|  | Sexual contact | 0.9 | 0.4 – 1.8 | 0.8 | 0.4 - 1.6 |
| **Sex at venues** | No | Ref. |  | Ref. |  |
|  | Yes | 0.9 | 0.6 – 1.2 | 0.9 | 0.7 - 1.3 |
| **Group sex** | No | Ref. |  | Ref. |  |
|  | Yes | 0.8 | 0.6 – 1.1 | 0.8 | 0.6 - 1.2 |
| **High-risk of MPXV exposure <sup>c</sup>** | No | Ref. |  | Ref. |  |
|  | Yes | 1.0 | 0.7 – 1.4 | 1.1 | 0.8 - 1.4 |
| **Number of sex activities** | 1-2 | Ref. |  | Ref. |  |
|  | 3+ | 1.5 | 1.0 – 2.3 | **1.5** | **1.0 - 2.1** |

PEP = post-exposure prophylaxis; MSM = men who (also) have sex with men; <sup>a</sup> Mild MPX= cases experiencing either 1-2 systemic symptoms (e.g. fever, lymphadenopathy) and/or skin lesions on one body location (e.g. head, limbs, trunk, peri-anal, genital) only. More severe MPX = cases experiencing ≥3 systemic symptoms and/or skin lesions on ≥2 body locations and/or hospitalization due to MPX; <sup>b</sup> adjusted for age-group; <sup>c</sup> including PrEP-HIV use; <sup>d</sup> either group sex, sex at venues/places or contact with MPX case.

### Smallpox vaccine effectiveness against more severe monkeypox

The crude VE of the first-generation smallpox vaccine against more severe MPX was estimated at 55% (95% CI 20%; 75%). After adjustment for age the VE remained similar (58%; 95% CI; 17%; 78%) (Table 5).

**Table 5.** Vaccine effectiveness of the first-generation smallpox vaccine against more severe Monkeypox, the Netherlands, May 20 to August 8 2022.

| Vaccination status | Number of mild MPX cases | Number of severe MPX cases | Total cases | Crude vaccine effectiveness | 95% CI | Adjusted vaccine effectiveness<sup>a</sup> | 95% CI |
| --- | --- | --- | --- | --- | --- | --- | --- |
| Unvaccinated | 28 | 63 | 91 | Ref. |  | Ref. |  |
| Vaccinated | 58 | 59 | 117 | 55% | 20%; 75% | 58% | 17%; 78% |

Mild MPX= cases experiencing either 1-2 systemic symptoms (e.g. fever, lymphadenopathy) and/or skin lesions on one body location (e.g. head, limbs, trunk, peri-anal, genital) only. More severe MPX = cases experiencing ≥3 systemic symptoms and/or skin lesions on ≥2 body locations and/or hospitalization due to MPX <sup>a</sup> adjusted for age in 44–50 , 51–55 and 56+ years.

## Discussion

We describe the first outbreak of MPX in the Netherlands, which is part of the international outbreak that started early May 2022 in Europe and extended globally.^2, 4^ The first 1000 MPX cases, reported between May 20 and August 8 2022, were predominantly MSM aged 31-45 years, which is similar to the outbreaks in other countries. ^1, 22-24^ The vast majority of infections were acquired through sexual contact with casual partners at family/home or recreational settings including LGBTQIA+ venues in the Netherlands. This indicates that, although some larger upsurges occurred from point-source and/or travel related events the outbreak is mainly characterised by sustained transmission within the Netherlands.

Most MPX cases reported both systemic symptoms and skin lesions, and for about half of the cases skin lesions developed up to 9 days (median 2 days) after systemic symptoms onset. This highlights the importance of testing for MPXV in persons who are at high risk of MPX even in absence of skin lesions initially, as well as clinical follow-up after testing positive. Delay in testing or ending self-isolation too soon enhances the risk of further transmission.

We found that more severe MPX was associated with having one or more comorbidities, and observed no mortalities. Previous international reports shows that MPX is more severe in elderly people, pregnant women, children and immunocompromised with mortality rates of 1-10%.^25, 26^ In the current outbreak, with ongoing sexual person-to-person transmission, few hospitalisations and deaths have been reported internationally, and it seems that less than 1/1000 cases die from MPX related complications. ^25-27^ This difference in morbidity and mortality could be explained by bias in population involved, as the current circulation is in a select group excluding children, pregnant women and with a low prevalence of immunocompromised; selection bias due to health system constraints in Africa with possibly only registration of severe cases; by differences in the mode of transmission, or the specific strain involved.

Additionally, we found that more severe MPX was associated with having participated in more (3+) different sexual activities. This result could indicate the importance of direct (sexual or close skin) contact instead of respiratory droplets as a mode of transmission in this outbreak with only few cases (4%) reporting coughing or other respiratory symptoms (8%), and no or little indication of indirect transmission.

Few studies have looked at the VE of the first-generation smallpox vaccine against (severe) MPX, and there is limited evidence on its immunogenicity against MPX. An observational study from 1988 showed an estimated VE of 85% against MPX disease among household contacts. ^28, 29^

We did not have adequate data to allow estimation of the VE of first-generation smallpox vaccines to protect against MPX infection. However, we found a VE against more severe MPX of 58% (95% CI 17-78%). This suggests moderate protection against severe MPX symptoms on top of any possible protection by the first-generation smallpox vaccine against MPXV infection and disease.

There are several factors that can influence VE estimates. Differences in timepoints of the first contact with the PHS between mild and more severe cases can bias VE estimates: e.g. mild cases might be contacted earlier by the PHS than the more severe cases, and symptoms that developed after that contact might not be registered. However, within our population for the VE analysis, there was no significant difference in time between date of onset and the first contact with the PHS between mild and more severe MPX cases: both mild and more severe MPX cases were contacted up to 16 days (median 6 days) after onset of first symptoms (p=0.58).

Furthermore, we used surveillance data with self-reported vaccination status to calculate VE, potentially leading to misclassification. However, this misclassification is unlikely to depend on the outcome and therefore more likely to lead to an underestimation of VE.

It is uncertain if our VE estimate is transferable to the third-generation smallpox vaccine (Imvanex®). In the Netherlands Imvanex® is used as PEP in high-risk contacts and is, since the end of July, also available as PrEP for populations at highest risk, to prevent MPX infection. VE estimates of Imvanex® against MPXV infection, MPX symptoms and/or duration of illness could impact public health control measures such as vaccination strategies and isolation.

Although the outbreak is still ongoing, the number of reported cases by date of symptom onset reached a peak in the first half of July 2022. This might be explained in part by fewer infected cases seeking care during the holiday season or concerns about isolation measures, or a positive effect of public health control measures such as heightened awareness in the at risk group, case finding, isolation of cases and notification of contacts, and acquired immunity due to natural infection or vaccination in persons with high-risk of MPXV exposure. The full extent of the outbreak, including potential asymptomatic MPXV infections, might have been higher. ^30^ Serological population studies and case-control studies might give such indication on the wider extent of exposure and infection.

## Conclusions

The current outbreak of MPXV among MSM is mainly characterised by sustained transmission through direct (sexual) contact. Individuals who received a first -generation smallpox vaccine have a lower probability of developing more severe MPX.

## Supporting information

Supplement 1 - Box 1 and 2

## Data Availability

All data produced in the present study are available upon reasonable request to the authors

