## Supplement 1 - Box 1 and 2 for "Monkeypox outbreak in the Netherlands in 2022: public health response, epidemiological and clinical characteristics of the first 1000 cases and protection of the first-generation smallpox vaccine"

**Authors and affiliations:**

Catharina E. van Ewijk ^1,2^, Fuminari Miura ^1,3^, Gini van Rijckevorsel ^1,4^, Henry J.C. de Vries ^5,6,7,8^, Matthijs R.A. Welkers ^4,9^ , Oda E. van den Berg ^1^, Ingrid H.M. Friesema ^1^ , Patrick van den Berg ^1^, Thomas Dalhuisen ^1^, Jacco Wallinga ^1,10^, Diederik Brandwagt ^1^, Brigitte A.G.L. van Cleef ^1,4^, Harry Vennema ^1^, Bettie Voordouw ^1^, Marion Koopmans ^11^, Annemiek A. van der Eijk ^11^, Corien M. Swaan^1^, Margreet J.M. te Wierik ^1^, Tjalling Leenstra ^1^, Eline Op de Coul * ^1^, Eelco Franz * ^1^, the Dutch Monkeypox Response Team

* shared last author

1. Centre for Infectious Disease Control, National Institute for Public Health and the Environment (RIVM), Bilthoven, the Netherlands
2. European Programme for Intervention Epidemiology Training (EPIET), European Centre for Disease Prevention and Control (ECDC), Solna, Sweden
3. Centre for Marine Environmental Studies, Ehime University, Matsuyama, Japan
4. Department of Infectious Diseases, Public Health Service of Amsterdam, the Netherlands
5. Amsterdam UMC location University of Amsterdam, Department of Dermatology, Amsterdam, the Netherlands
6. Amsterdam Institute for Infection and Immunology, Infectious Diseases, Amsterdam, the Netherlands
7. Centre for Sexual Health, Department of Infectious Diseases, Public Health Service of Amsterdam, the Netherlands
8. Amsterdam Institute for Global Health and Development, Amsterdam, the Netherlands
9. Amsterdam UMC location AMC, University of Amsterdam, Department of Medical
    Microbiology and Infection Prevention, Amsterdam, the Netherlands
10. Department of Biomedical Data Sciences, Leiden University Medical Centre, Leiden, the Netherlands
11. Department of Viroscience, Erasmus Medical Centre, Rotterdam, the Netherlands

**Dutch Monkeypox Response Team:**  Birgit van Benthem, Diederik Brandwagt, Annemiek A. van der Eijk, Hanna Bos, Colette van Bokhoven-Rombouts, Lian Bovée, Chantal P. Bleeker-Rovers, Brigitte van Cleef, Alje P. van Dam, Rik van Dael, Jaap van Dissel, Pauline Ellerbroek, Catharina van Ewijk, Eelco Franz, Corine GeurtsvanKessel, Joke van der Giessen, Hannelore Götz, Josette Häger, Susan van den Hof, Elske Hoornenborg, Putri Hintaran, Jorgen de Jonge, Rosa Joosten, Marion Koopmans, Kevin Kosterman, Jente Lange, Tjalling Leenstra, André J. Meeske, Liesbeth Mollema, Eline Op de Coul, Demi Reurings, Gini van Rijckevorsel, Helma Ruijs, Corien M. Swaan, Linda Smid, George Sips, Sacha F. de Stoppelaar, Albert Vollaard, Bettie Voordouw, Harry Vennema, Henry J.C. de Vries, Karin Ellen Veldkamp, Klaartje Weijdema, Geert Westerhuis, Margreet J.M. te Wierik, Matthijs Welkers, Toos Waegemaekers, Jacco Wallinga, Paul Zantkuijl.

**Corresponding author:**

Catharina van Ewijk, Centre for Infectious Disease Control Netherlands, National Institute for Public Health and the Environment (RIVM), Antonie van Leeuwenhoeklaan 9, 3721 MA Bilthoven, the Netherlands

**Abstract**

In early May 2022 a global outbreak of monkeypox (MPX) started among persons without a travel history to regions known to be enzootic for monkeypox-virus. On August 8 2022, the Netherlands reported its 1000^th^ monkeypox case representing a cumulative incidence of 55 per million population, one of the highest cumulative incidences worldwide. Here we describe the epidemiological characteristics and clinical presentation of the first 1000 monkeypox cases in the Netherlands, within the context of the public health response. Additionally, we explored risk factors for and estimated the protective effect of first-generation smallpox vaccine against more severe MPX.

The first 1000 MPX cases, reported between May 20 and August 8 2022, were predominantly MSM aged 31-45 years. The vast majority of infections were acquired through sexual contact with casual partners in private or recreational settings including LGBTQIA+ venues in the Netherlands. This indicates that, although some larger upsurges occurred from point-source and/or travel related events, the outbreak is mainly characterised by sustained transmission within the Netherlands. More severe MPX was associated with having one or more comorbidities as well as having participated in more (3+) different sexual activities 21 days before symptom onset. We found a vaccine effectiveness of the prior first-generation smallpox vaccine against more severe MPX of 58% (95% CI 17-78%), suggesting moderate protection against more severe MPX symptoms on top of any possible protection by this vaccine against MPXV infection and disease.

**Supplement 1**

**Confirmed case**

A person with a laboratory confirmed MPXV (PCR positive for Orthopoxvirus with or without additional MPXV confirmation by sequencing or MPXV-specific PCR).

**Probable case**

A person with skin lesions consistent with monkeypox on (a part of) the body with symptom onset after March 1, 2022, and/or a person with complaints consistent with proctitis (including anal pain) in which the complaints occurred after March 1 2022, and optionally one or more systemic symptoms * consistent with monkeypox

AND

one or more of the following criteria:

1) Contact with a confirmed or probable case of monkeypox 21 days prior to symptom onset.

2) A man who (also) has sex with men.

3) A female partner of a man who (also) has sex with men.

4) A person (regardless of sexual orientation) who indicates having had multiple sexual contacts, whether anonymous or not, or paid for (e.g. at sex parties) 21 days before symptom onset.

**Possible case**

A person with skin lesions consistent with monkeypox on (a part of) the body with symptom onset after March 1, 2022, and/or a person with complaints consistent with proctitis (including anal pain) in which the complaints occurred after March 1 2022, and optionally one or more systemic symptoms* consistent with monkeypox

AND

without an epidemiological link with a person who has clinically suspected or confirmed varicella, and an infection with another known causative agent of a similar skin appearance, such as herpes zoster, (primary) herpes simplex, primary or secondary syphilis, is considered unlikely.

* Fever (>38.5°C), headache, myalgia, backpain, malaise, (usually painful) lymphadenopathy (localized or generalized).

**Box 1 - Clinical and epidemiological criteria for classification of monkeypox cases, the Netherlands, as of 8 August 2022.**

**Box 2 - Categorisation of high medium and low risk contact of monkeypox cases during the outbreak in The Netherlands, as of 8 August 2022.**

**High risk contact:** a person with (one) of the following type of contact with a monkeypox case during their infectious period

a. Any type of sexual contact
b. Intensive skin-skin contact (such as hugging, kissing)
c. Household contact, excluding intensive skin-skin contact and sexual contact
d. Unprotected direct contact with a monkeypox patient and/or contaminated patient material
e. Laboratory employees with unprotected exposure accident to contaminated material

**Medium risk contact:** a person with (one) of the following type of contact with a monkeypox case during their infectious period

a. Unprotected prolonged (cumulative more than 2 hour) face-to-face contact within 1.5 meter distance (such as caregivers without a mouth and nose mask, in social situations, including public transport)

**Low risk contact :** a person with (one) of the following type of contact with a monkeypox case during their infectious period

a. Unprotected short (cumulative less than 2 hour) face-to-face contact within 1.5 meter distance (such as caregivers without PPE)
b. Fellow travelers with a journey time (more than 8 hours) within 1.5 meter distance (1-2 seats around the index)
c. Social contact short (cumulative less than 2 hour) face-to-face contact within 1.5 meter distance

**No risk:** a person with (one) of the following type of contact with a monkeypox case during their infectious period

a. Caregivers (including lab staff) with full PPE: direct contact with a monkeypox patient and/or contaminated patient material
b. Caregivers and social contacts with unprotected exposure at more than 1.5 meter distance (regardless of duration)
